# Position: a study protocol for the prevention of fall injuries in French Special Forces selection courses using a body-centered intervention

**DOI:** 10.1101/2023.07.13.23292623

**Authors:** Loucas Obligi, Mathieu Bertrand, Mathieu Boivent, Simon-Pierre Corcostegui, Pierre-Emmanuel Coz, Clément Derkenne, Vincent Des Robert, Victor Hurpin, Jauffrey Hus, Benoît L’Hermitte, Laurent Lely, Edouard Patey, Emeric Romary, Luc Saint-Jean, Alexandre Trente, Marine Turpin, Nicolas Vertu, Charles Verdonk, Anaïs M. Duffaud

## Abstract

**Introduction:** The Military Physical and Sports Training program was developed by the French Army in order to train, optimize, and maintain individual readiness. Although the health benefits of sport practice do not need to be demonstrated, such activities can cause acute musculoskeletal injuries that need to be addressed. The prevalence of lower limb injury is rather high in the French military population and, in particular, ranges from 15 to 45% during Special Forces selection courses. Thus, this project aims to investigate the efficiency of a body-centered program designed to enhance body awareness. The program seeks to train the mind to actively pay attention to body information, while the latter is viewed as a protective factor against fall injuries. We assume: (i) that postural control can be improved by enhancing the level of body awareness; and (ii) that greater postural awareness could be beneficial in reducing the risk of fall injuries. The body-centered prevention program is based on the Optimization of the Resources of the Armed Forces (ORAF) intervention, which focuses on mental preparation and recovery, and has been deployed in the French Army for many years

**Method and analyses:** The study focuses on five French Special Forces selection courses (400 soldiers/ participants). It is divided into two stages (year 1, year 2). The first year is dedicated to data collection from the control group (200 participants), while in the second year the ORAF intervention will be deployed. The main objective is to evaluate the effectiveness of the ORAF intervention in reducing the rate of fall injuries during military selection, based on a multidisciplinary method that captures demographic, biological, biometric, clinical, and para clinical measures.

**Registration number:** IDRCB number 2021-A02108-33, Clinical Trial: NCT 05451394

## Introduction

In military settings, physical activity and sports play an important role in developing and optimizing readiness. The Military Physical and Sports Training program, run by the French Army, aims to enable all military personnel to acquire the physical and mental skills required for full proficiency. Moreover, maintaining operational capacity throughout a military career relies on, among other things, the health benefits of sports practice, particularly with regard to the prevention of stress-related chronic disease (cardiovascular disease, depression, etc.) [1, 2]. However, physical activity and sport can also cause both acute injuries, due to a fall or twist (such as sprains or fractures) and chronic pathologies resulting from overtraining/ overuse (such as tendinopathies). In this context, the prevalence of lower limb injuries incurred during physical activities in the French military population is high – not only among enlisted soldiers during their first year of service (15–20%) [3, 4] but also in operational units, or during special operations’ selection courses (30–45%) [5–9]. Beyond their consequences for the soldier’s health, these injuries negatively impact the operational capacity of the country’s armed forces (due to disability discharge) and incur a non-negligible economic cost [10]. Thus, it appears that a prevention program designed to prevent, or at least reduce the risk of such injuries would be of interest to improve the readiness of the French Army. In the context of evidence based prevention, we need to identify risk factors that could be modified in order to reduce their influence on the injury rate. From this perspective, the present project aims to investigate a cognitive mechanism—body awareness—as a risk factor for fall injuries, and as a target for a prevention strategy.

Several (published and unpublished) epidemiological studies have been conducted in selected military contexts, notably during the selection process to join Special Forces units. These studies highlight the high prevalence of acute traumatic injuries due to falls, resulting in candidates failing the selection course [5, 8, 9, 11]. The failure rate for selection courses run for French Special Forces is about 90%, and 70% of failures are due to medical reasons. Specifically, 60% of medical diagnoses fall into the category of acute traumatic injuries, and 87% of the latter are identified as resulting from falls due to a loss of balance. The high prevalence of fall injuries suggests that functional/ dysfunctional postural balance could play a central role in an individual’s ability to be successful in the physical activities that characterize Special Forces selection courses.

In practice, the physical activities that participants engage in make significant demands on postural balance: trainees carry heavy loads (a combat bag and their weapon, weighing around 15 kg) for prolonged periods of time, and complete extreme challenges that are run at height, regardless of weather conditions. Furthermore, it is important to emphasize that the activities carried out by military units are highly intensive, both physically and psychologically. Trainees are continuously exposed to high-intensity stressors (e.g., lack of sleep/ sleep deprivation, a wet environment, uncertainty regarding what the day will hold/ the planning of activities, time pressure). The latter challenges are combined with demanding physical exercises (performed in combat gear) that aim to train them to effectively cope with the stressful situations that characterize Special Forces operations [5]. The activities that make up the selection course are thought to activate a psycho-neurobiological stress response that induces changes at psychological, neural (e.g., activation of the amygdala), and biological (e.g., cortisol release) levels. For a comprehensive review of neurobiological aspects of stress see [12]. The neurobiological mechanisms that underpin a stress response are classically viewed as an aspecific (i.e., the stress response is similar across psychological or physical stressors), and adaptive strategy. This process enables the organism to respond to changes in its environment that affect its integrity, to adapt to these changes if they persist, and to adapt to any metabolic needs resulting from changes in the internal (body) or the external (environment) milieu. The stress response was originally described as ‘general adaptation syndrome’ [13].

### Body awareness: a candidate cognitive target for sports injury prevention programs

Several neurophysiological systems contribute to postural balance. The latter is not just a muscular reflex, as it involves different high-level cognitive processes such as attention, visuo spatial perception, and executive functions [14]. Sensory information from peripheral receptors (located in visual, vestibular, and somato-sensory systems) is integrated at the level of the central nervous system, which then controls muscles that orient the different parts of the body with respect to gravity [15]. The cognitive mechanisms associated with postural body awareness refer to the attention the individual gives to information that is derived from somato sensory and vestibular systems [16]. The more attention individuals give to their body, the more sensory information they have available to control their posture (thus preventing a fall). In other words, greater postural control could be supported by a higher level of body awareness. In the present study, we assume that the individual level of postural awareness may influence the risk of fall injuries. Specifically, we hypothesize that a high level of postural awareness may contribute to preventing fall injuries due to a higher level of postural control (Fig. 1).

**Fig. 1.**
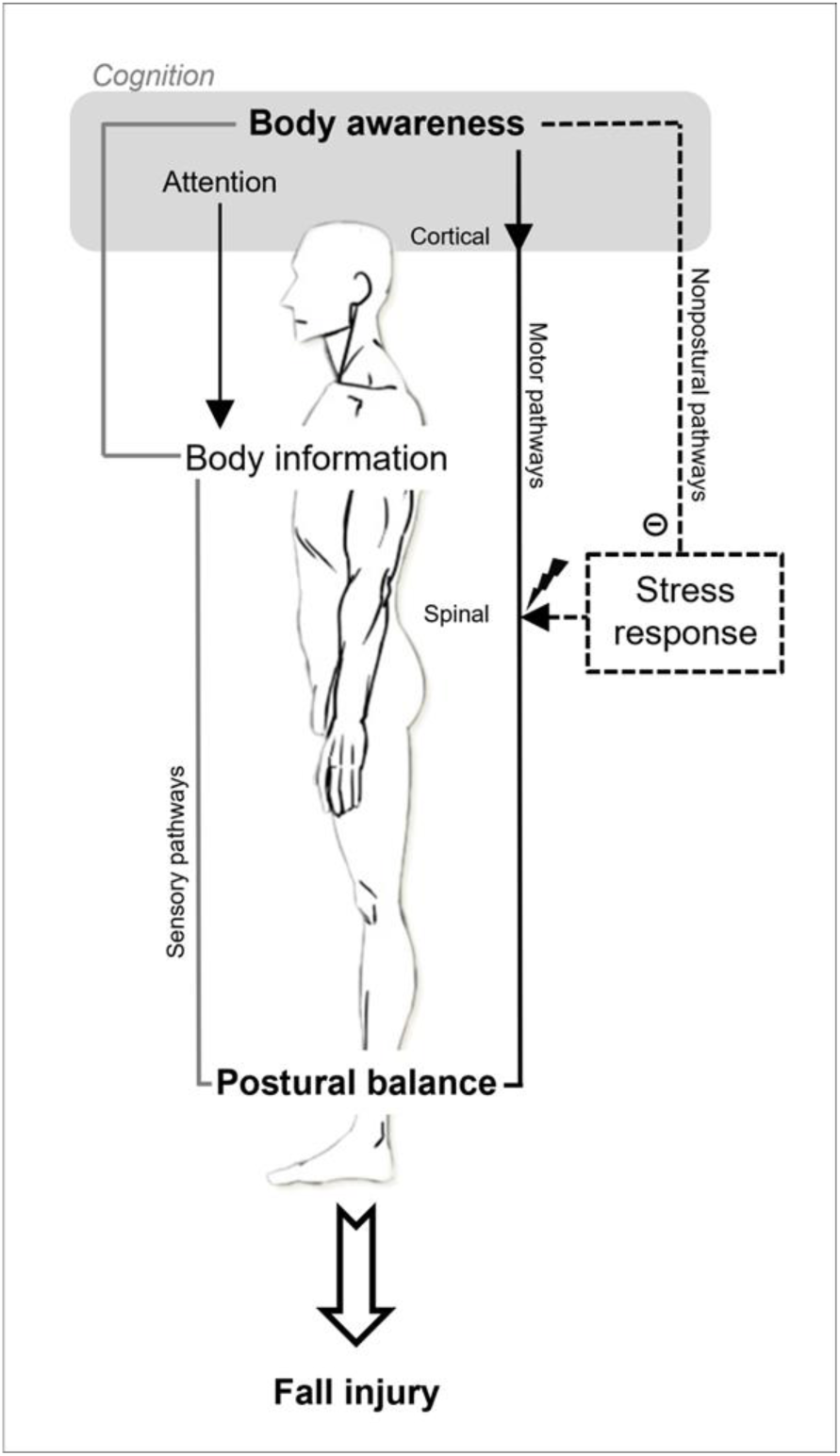
Schematic illustration of our cognitive approach to postural balance. The illustration depicts the relation to fall injury in physically demanding, high-stress situations (such as a Special Forces selection course). The internal representation of the body position is the result of the integration, at the cortical level, of multisensory information from peripheral sensors (the gray/ black line). The higher-level cognitive mechanism governing body awareness involves active (i.e., top-down) attention to body information, and influences postural balance either: (i) directly, by acting on descending motor pathways (the solid black line), as a central component of the sensorimotor pathways forming the postural control loop; or (ii) indirectly, by down-regulation of non-postural pathways (the dashed black line), such as the stress response, which may alter the postural control loop at the spinal level (Source: image of body from www.dessindigo.com).

Furthermore, it should be noted that the neurophysiological systems involved in postural control also receive inputs from brain structures outside the sensory pathways. Thus, for example, the degree of excitability of the postural myotatic reflex at the level of the spinal cord is modulated by subcortical structures (notably the periaqueductal gray matter, and the amygdala) that are activated during the stress response [17]. In addition, body awareness seems to play an important role in the regulation of the stress response, by contributing to the identification, evaluation, and regulation of the body’s internal physiological state [18]. Altogether, this suggests that the body awareness mechanism could influence the occurrence of fall injuries in two ways: either (i) it is a central and direct player in the sensorimotor pathways governing postural control; and/ or (ii) it acts indirectly (e.g. via the stress response) on the non-postural pathways that affect posture (Fig. 1).

### Optimization of the resources of the armed forces

The present project focuses on the cognitive mechanism of body awareness as a risk factor for fall injuries during military physical activities. In this context, it is essential to assess the modifiability of the risk factor, in order to develop an injury prevention strategy. *Modifiability* means that the risk factor is amenable to an intervention that will control its influence on the occurrence of the injury [19]. The project aims to test the effect of the ORAF (optimization of the resources of the armed forces) method on the cognitive mechanism of body awareness. ORAF refers to a form of mental preparation, where the aim is to learn a set of techniques and strategies that enable an individual to mobilize his or her physical and psychological resources to the best of their ability, as a function of the demands of the situation. The techniques were created in the 1990s by Dr. Edith Perreault-Pierre to meet needs that had never been addressed before in the armed forces: mental preparation and recovery [20].

Initially developed for the Air Force, and focused on stress management, they take into account the human factor (in the context of flight safety), and are now being deployed in all other army corps in France. They have recently been enriched with six new techniques that seek to respond as best as possible to the demands of the field and, if necessary, high intensity engagements. They are based on basic procedures that are widely used in top-level sport. The method takes the form of a customizable ‘toolbox’ that is expected to enable trained personnel to carry out their missions successfully, while preventing the deleterious effects of stress. The literature contains many reports of the beneficial effects of mental imagery, internal dialogue, relaxation and mindfulness techniques on performance, health and well-being [21, 22]. Similarly, ORAF studies conducted in a military population have mainly focused on sleep, stress, and psycho-cognitive and physical performance [23–27]. Although the results of these studies are difficult to compare (there are several before-and-after interventional studies, and one randomized controlled trial), taken together, the findings provide a body of evidence that supports the clinical and operational benefits of ORAF.

Within the context of our protocol, the ORAF program focuses on tools and techniques that are expected to specifically enhance postural awareness (i.e., body-centered training).

### Hypotheses

The rationale for our hypotheses relies on a cognitive approach to postural balance, and its relation to fall injuries, as described above (see Fig. 1). We assume that a low level of postural body awareness is a risk factor for fall injuries during French Special Forces selection courses, due to impaired postural control. Our first hypothesis is that a body-centered prevention program, specifically an intervention that enhances body awareness by training the mind to actively pay attention to the body, will be beneficial in reducing the risk of fall injuries. Therefore, we expect that the incidence of fall injuries will be significantly lower in the ORAF group compared to the active control group. Secondly, we hypothesize that the risk of fall injuries can be predicted from an analysis of the postural signal, recorded prior to the beginning of the selection course using posturography. We expect that a numerical model can successfully predict fall injury status at the individual level from the postural signal, with accuracy of over 50%.

## Methods

### Objectives

The primary objective is to evaluate the effectiveness of the ORAF intervention in preventing fall injuries during military selection courses, within the four weeks following the inclusion visit.

The following four secondary objectives relate to medical and mechanistic/ scientific purposes:

i. To test the hypothesis that the ORAF intervention will reduce the risk of fall injuries by increasing postural awareness and modulating postural balance, as measured by the self-report Postural Awareness Scale (PAS) and posturography, respectively. Here, fall injury refers to acute injuries such as ankle sprain, bone fracture, wounds, or contusions that result from a fall.
ii. To test the hypothesis that the ORAF intervention will reduce the risk of fall injuries by decreasing the intensity of the psychobiological stress response.
iii. To develop a predictive model of fall injuries occurring during the selection course from clinical and paraclinical measures of postural balance, including the Balance Error Scoring System (BESS) test and posturography, assessed at the initial visit before the selection course begins (see below).
iv. To evaluate the persistence over time of the effectiveness of an ORAF intervention in preventing the risk of fall injuries.

### Experimental design and setting

This study is being conducted in five French Special Forces units (the National Gendarmerie, the 1st Marine Infantry Parachute Regiment, the 13th Parachute Dragoon Regiment, the 10th Air Parachute Commando, and the Fusiliers Marins [Navy Riflemen] School). It will take place during selection courses that are organized independently in each of the units (courses are run at different times in the year, and have different durations). Participation will last up to four weeks (if he/ she does not leave the selection process before the completion of the study visit). The whole project is planned to last two years (two interventions will be run over two years within the five units).

### Randomization

The intervention (ORAF versus the active control) is not randomized at the participant level in order to avoid contamination bias (a participant being inadvertently exposed to the other intervention). As the selection courses are carried out in crowded conditions, the risk of inter participant influence between the ORAF and the active control program is quite high. We will therefore run the study over two successive years (Y and Y+1). During the first year (Y) participants will take part in active control sessions, and during the second year (Y+1) a second cohort will benefit from the ORAF intervention. It should be noted that there is no randomization of the order of the two interventions between units; this is because if a beneficial effect of the ORAF intervention is observed in year Y, it would be very difficult to get agreement from the different units to implement the active control program in year Y+1.

### Eligibility criteria

Participants will have to meet the following criteria:

Inclusion criteria:

Signing an informed consent form to participate in the study
Involved in a selection course
Man or woman aged over 18 years
Covered by the French National Health Service

Exclusion criteria:

Participants with previous advanced ORAF training (> 10 hours)
Refusal to participate
Persons covered by articles L1121-5 to L1121-8 of the Public Health Code

### Participant timeline (Fig. 2)

**Fig 2.**
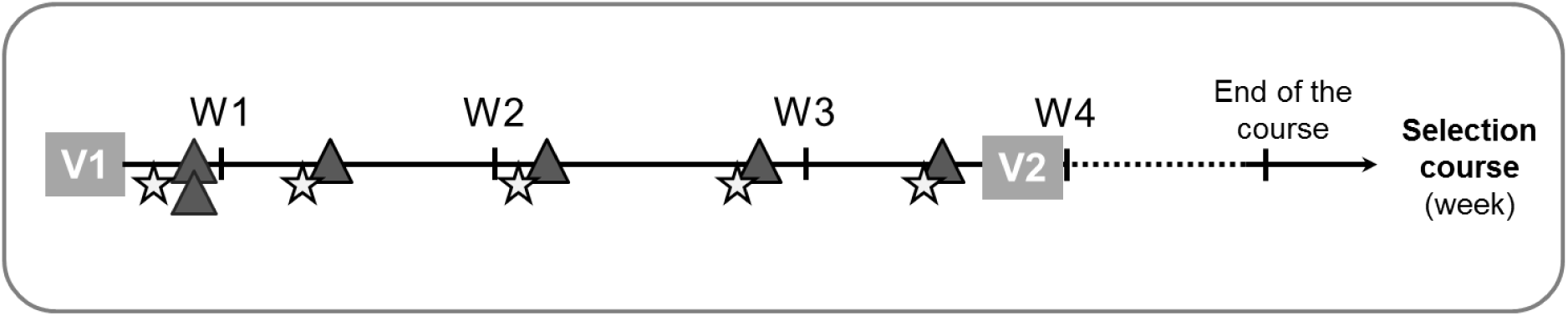
Timeline for each study participant (in weeks). V1: inclusion visit (saliva sampling, posturography, and the BESS). V2: end-of-study visit (saliva sampling, and posturography). Gray triangle: a one-hour session, either during the active control program (first year), or the ORAF intervention (second year). Star: Self-administeed questionnaires (PAS, PSS, fatigue).

### Inclusion visit

Prior to inclusion, a general overview of the study will be presented to all candidates, and they will be given time to consider whether they would like to participate.

The inclusion visit will start with the signature of an informed consent form, then the initial assessment begins. Thereafter, we will collect individual demographic and biometric data (age, gender, height, and weight). Posturography measurements include the recording of the postural signal using the FEETEST 6 platform (TECHNO CONCEPT^®^, France). During posturography, participants are instructed to stand quietly on the platform with their eyes closed (arms by their sides and head in a normal, forward-facing position). Due to the technical specifications of the FEETEST 6 platform, the recording will last 52 seconds [28–30]. In addition, they will be asked to take the Balance Error Scoring System (BESS) clinical test, and a 4 ml saliva sample will be collected.

Participants will subsequently complete socio-demographic and military self-administered questionnaires, along with postural awareness (PAS), perceived stress (PSS), and fatigue (using a visual analogic scale, ranging from 0 to 10) self-administered questionnaires. Altogether, this visit will last approximately 20–30 minutes per participant.

### End-of-study visit

The subject’s participation in the study will end in one of two ways:

i. either on the day of the final visit; or
ii. on the day of discharge from the selection course for whatever reason (e.g., injury, personal reasons, or the instructor’s decision) if this occurs before the end of the intervention (either ORAF or active control).

In all cases, posturography measurements will be recorded, a 4 ml saliva sample will be collected, and self-administered questionnaires (PAS, PSS, and fatigue) will be completed. This visit will last approximately 20 minutes per participant.

Thus, each participant will attend a minimum of two sessions. However, there may be additional visits, notably if a subject needs a medical consultation following a fall injury. This visit will take place on the day the trauma occurs. A doctor (or a member of the medical staff) will collect medical information related to the trauma, and perform a posturography measurement. In addition, the participant will complete PAS and PSS questionnaires.

Any adverse events, and other unintended effects of the study will be reported in compliance with the French public health code.

### Programs

The intervention program (either active control or ORAF) is divided into six, one-hour sessions spread over three weeks. The study will run over two successive years (Y and Y+1). During the first year (Y), participants will take part in active control sessions, and during the second year (Y+1) a second cohort will benefit from the ORAF intervention. Details of the programs’ contents are provided in Supplementary File 1.

The ORAF intervention will consist of practical training given by a qualified instructor within each unit. The instructor is a member of the military who has completed the ORAF instructor training course delivered by the National Defense Sports Center. The intervention will include theoretical instruction and practice related to ORAF techniques identified as relevant to improve body awareness. The content of the six sessions making up the program has been defined and validated by ORAF instructors in the five units, under the supervision of another ORAF instructor who is coordinating the intervention.

The active control program will be conducted in the same way. These sessions will address high-level cognitive processes such as memory, reasoning, or creativity. Each session will be divided into two parts: the first, theoretical part describes the processes that are the focus of the session, then the second part focuses on practical exercises in relevant situations. Whenever possible, the same ORAF instructor will be in charge of running the six sessions over the two years of the study (both ORAF and active control programs).

### Endpoints

The primary endpoint is the incidence of fall injuries due to postural imbalance in each of the two groups (ORAF *vs* active control). ‘Fall injury’ is defined in our protocol as an acute, traumatic injury that is suspected to be caused by postural imbalance, based on a clinical examination (an interview with the patient, and a physical examination of the injury) by the medical team supporting the selection course.

There are four secondary objectives. The first has two endpoints (secondary-i in the table 1): 1) a comparison of PAS scores pre- and post-intervention, and at the beginning of each session; and 2) change in postural balance measurements pre- and post-intervention (measured with posturography).

**Table 1.**
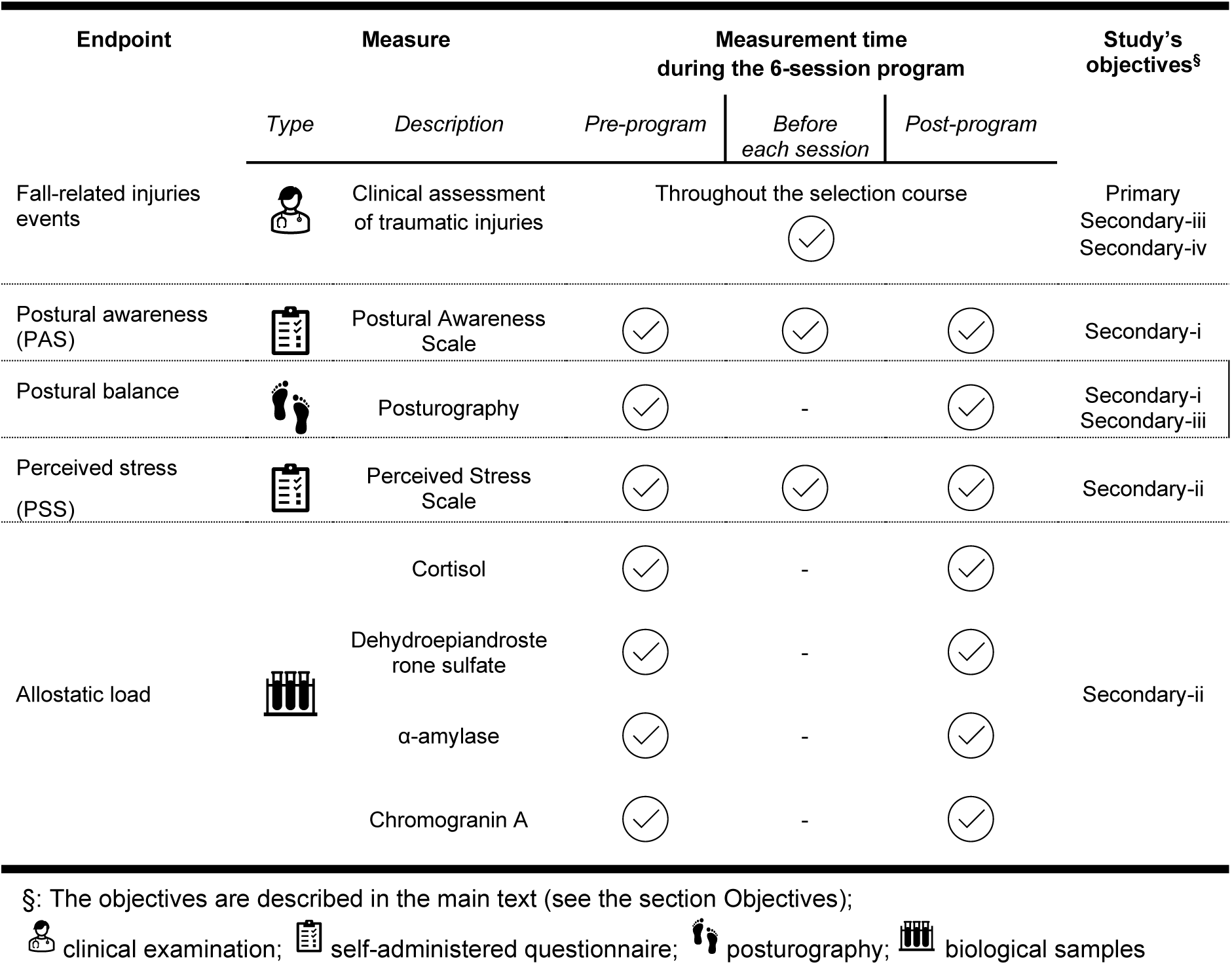
Endpoints of the protocol.

| Endpoint | Measure |  | Measurement time during the 6-session program |  |  | Study's objectives<sup>§</sup> |
| --- | --- | --- | --- | --- | --- | --- |
|  | Type | Description | Pre-program | Before each session | Post-program |  |
| Fall-related injuries events |  | Clinical assessment of traumatic injuries | Throughout the selection course |  |  | Primary<br>Secondary-iii<br>Secondary-iv |
| Postural awareness (PAS) |  | Postural Awareness Scale |  |  |  | Secondary-i |
| Postural balance |  | Posturography |  | - |  | Secondary-i<br>Secondary-iii |
| Perceived stress (PSS) |  | Perceived Stress Scale |  |  |  | Secondary-ii |
| Allostatic load |  | Cortisol |  | - |  | Secondary-ii |
|  |  | Dehydroepiandrosterone sulfate |  | - |  |  |
| | | α-amylase | | - | | |
|  |  | Chromogranin A |  | - |  |  |

§: The objectives are described in the main text (see the section Objectives);
clinical examination; self-administered questionnaire; posturography; biological samples

The second also has two endpoints (secondary-ii in the table 1): 1) a comparison of PSS scores pre- and post-intervention, and at the beginning of each session; and 2) change in allostatic load in saliva pre- and post-intervention (the corticotropic axis: cortisol and dehydroepiandrosterone sulfate; the autonomic nervous system: α-amylase and chromogranin A).

The endpoint of the third secondary (secondary-iii in the table 1) objective is the predictive accuracy of the risk of fall injuries using our machine learning-based algorithm.

Finally, the endpoint of the fourth secondary objective (secondary-iv in the table 1) is the incidence of fall injuries due to imbalance in each of the two groups (ORAF *vs* active control), between the end-of-study visit, and the end of the selection course.

Table 1 summarizes the different endpoints of the protocol.

## Materials

### Biological samples

Saliva sampling will be conducted according to the standard operating procedures described by the manufacturer. Each sample will be locally conditioned (5 x 500µl Eppendorf) and stored at −20°C. All biological samples will then be transferred to the IRBA laboratory, and stored at −80°C until analysis.

### Questionnaires

#### Socio-demographic and military data

Socio-demographic data include age, gender, hobbies, educational level, marital status, the practice of body-oriented activities (yoga, martial arts, etc.) and training modalities when preparing for the selection course. Regarding military data, the year of joining the army, military rank and specialty, level of ORAF training, and the number and type of military deployments will be collected.

#### The Postural Awareness Scale (PAS)

The 12-item Postural Awareness Scale measures two facets of postural body awareness: 1) *Ease/ familiarity with postural awareness* (PAS-EwPA), understood as effortless awareness of the body’s posture; 2) *Need for attention regulation with postural awareness* (PAS-NfA): in this case, awareness of the posture requires efforts to balance conscious cognitive processes and bodily needs. These two facets can be interpreted as opposite ends of a continuum of effort that is necessary to become aware of one’s posture. The questionnaire is scored using a seven-point scale, with responses ranging from 1 (not like me at all) to 7 (completely like me). For each of the two subscales, scores are calculated by summing the rating for all items; items related to the subscale *Need for attention regulation with postural awareness* (items 1, 2, 3, 4, 5 and 12) are reversed [31, 32].

#### The Perceived Stress Scale (PSS)

This questionnaire estimates how individuals evaluate their perception of stress. It consists of 14 items, which are rated on a 5-point Likert scale ranging from 1 (never) to 5 (often). A high score indicates a high level of perceived stress. A score above 27 is associated with a risk of psychopathology [33].

## Statistics

### Determination of sample size

To the best of our knowledge, no data on the effectiveness of the ORAF method in preventing sports injuries are available in the literature. However, discussions with military commanders and military doctors suggest that a reduction in the injury rate of about 15% would be clinically relevant. Preliminary epidemiological data from the five units indicate that the incidence rate of fall injuries is currently about 50%. Based on these findings, our sample size was determined *a priori* using GPOWER software (version 3.1.9.2) [34] with the following parameters: Statistical test: Fisher’s exact test; Tails: two; Proportion of injuries in the active control group: .50; Proportion of injuries in the ORAF group: .35; α error probability: .05; Power: .80. This resulted in *n* = 183 participants per group, and we plan to recruit 200 participants per group in order to compensate for potential drop-outs (estimated at 10%).

### Missing data

For missing data related to questionnaires and biological samples, we plan to use average imputation at the group level (ORAF or active control). Regarding physiological data related to the posturography, we plan to impute missing values using the *missForest* package, which is part of R [35].

### Analysis of the primary outcome

A comparative survival analysis will be performed to assess the effect of the intervention (ORAF *vs* active control) on the rate of fall injuries. Specifically, the Mantel–Cox log-rank test will be run to determine significant differences between the two interventions, using *survival*, which is another R package. We will conduct an intent-to-treat analysis to account for the potential bias introduced by participants withdrawing from the selection course for medical or non-medical reasons.

### General design

A general description of the study population will be presented for each group. For categorical variables, this will consist of the number of participants, and the percentage for each group. Central tendency and the variability of continuous variables will be described depending on whether or not the data follow a normal distribution (using the Shapiro–Wilk test): if there is a normal distribution, data will be summarized using means and standard deviations, if not, medians and ranges will be used.

### Analyses of secondary outcomes

For the first two secondary objectives, we will assess the effect over time of the ORAF intervention on self-reported postural awareness (PAS), postural features measured with posturography, self-reported measures of perceived stress (PSS), and biological measures of stress (cortisol, dehydroepiandrosterone sulfate, α-amylase, and chromogranin A). The analysis will implement a standard, repeated measures ANOVA and its Bayesian equivalent to extend insight, and guide our interpretation of significance (*p* values), and determine the likelihood of the alternative versus the null hypothesis [36]. Statistical analyses will be performed using JASP (version 0.13, https://jasp-stats.org/). Bayesian analyses will use default JASP priors (*r* scale fixed effects of 0.5, *r* scale random effects of 1, and *r* scale covariates of 0.354), and our model will be compared to the null model. For standard *post hoc* tests, we will apply the Holm correction for multiple comparisons [37]. Standard and Bayesian analyses will be performed with the assessment timepoint as a within-participants factor.

For the third secondary objective, we will follow a well-established machine learning methodology to design a classification model that assigns each participant to either “Fall” or “Non-fall” classes, based on his/ her postural measures, including the BESS test score and variables extracted from the posturography signal recorded during the initial visit. The machine learning pipeline will be implemented using MATLAB 2020b (The Mathworks^®^), including the Statistics and Machine Learning toolbox, the Deep Learning toolbox, and custom scripts. Our machine learning approach will comprise variable selection [38], specifically the random probe method [39], and the design of a neural network model [40]. The performance of the model will be assessed based on its predictive accuracy, which corresponds to the percentage of participants that are correctly classified (Equation (1)):

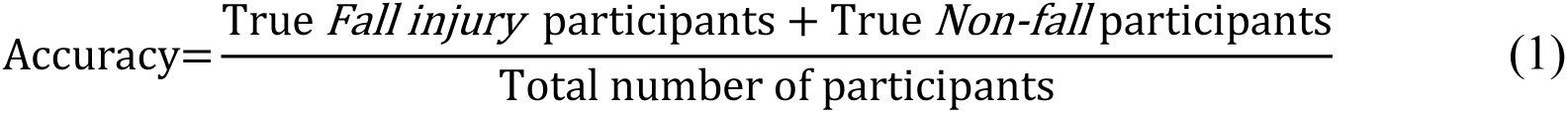

The fourth secondary objective will be analyzed by studying the persistence over time of the effectiveness of the ORAF intervention in preventing the risk of fall injuries, based on a comparative survival analysis (the Cox model). The survival curves of the two groups will be compared using the log-rank test. Here, the incidence of fall injuries will be recorded until the end of the selection course (from 8 to 12 weeks depending on the unit).

### Data collection and management

The clinical research associate coordinator of the *Direction centrale du service de santé des armées* will implement and conduct the study, according to the monitoring plan. As far as possible, she/ he will ensure the quality of data collection in each center in order to minimize missing data.

Personal information, data entry, coding, security, and storage will be processed in compliance with French legislation (the Act of 6 January 1978 on Data Processing, Data Files and Individual Liberties).

### Trial status

The study is currently recruiting participants.

### Ethics

Ethical approval has been obtained from the Sud-Est I Ethics Committee (reference n° 2020-114 of 13/09/2021), and the study will be conducted in accordance with ethical standards given in the 1964 Helsinki Declaration and its later amendments.

## Discussion

The present project aims to investigate cognitive body awareness as a causal mechanism in fall injuries during the physical activities that make up French Special Forces selection courses. The first aim is to examine whether a body-centered prevention program that seeks to improve body awareness by training the mind to actively pay attention to physical signals is more beneficial than an active control intervention in reducing the risk of fall injuries. A second goal is to identify analytical measures (physiology, self-reported and chemical) that best contribute to predicting fall injuries. The final endpoint will be to identify a set of assessments that could be turned into clinical tools to help military physicians predict the risk of fall injuries for each soldier, and develop a prevention program that could be deployed at group or individual level to reduce injury risk, thus, ultimately, reducing the rate of attrition in Special Forces selection courses.

Although previous research has revealed a large number of risk factors for sports injuries [19, 41], cognitive factors remain little-known, and poorly studied. To the best of our knowledge, the proposed study is the first to investigate body awareness as a cognitive factor influencing the risk of injury during physical activities. The literature proposes many theories and protocols that aim to limit the risk of injury; however, they tend to focus on physical exercises (Nordic Hamstring, Copenhagen Adduction, etc.) [42]. It appears from this literature that while some of these practices are applied, few have performance objectives [43].

In sum, this study provides additional support for the idea that mindfulness can be used to limit physical injury, notably in a context that makes high physical demands on the individual, and requires sustained performance. We can draw a parallel between this military requirement, and the quest for performance of the high-level athlete. The ORAF training program, as we note above, could therefore be seen as a way to prevent injury, and help the individual to become more self-aware of his or her practice. Moreover, it is interesting to note that while the literature shows that body awareness plays a critical role in many different health conditions, findings regarding its influence on the risk of mental disorders remain limited [44].

From a methodological perspective, our study relies on a naturalistic protocol. Measures and interventions are implemented during Special Forces selection courses, thus providing us with a unique opportunity to address our research questions using an ecologically-valid approach, and real-world scenarios. However, the drawback of such a protocol, which marks a shift away from the controlled laboratory setting, is that variability may affect the implementation of measures and interventions, notably due to the stringent operational constraints that are associated with Special Forces selection courses. For example, the frequency of the sessions run during the intervention will have to be adapted to each unit’s schedule, and may slightly differ (from two to four days). Similarly, the end-of-study visit may have to be scheduled at different times after the last session (from two to seven days). Finally, the intervention will be implemented during the selection course, and will include a relatively small number of sessions. It could be argued that implementing the prevention program before the selection course could increase its effectiveness in reducing injury risk. This is not possible due to the operational constraints faced by participants. Finally, it should be noted that the prevention program was designed by ORAF experts, and that the latter consider that a six-hour intervention is sufficient to improve the body awareness skills of participants, and contribute to reducing the risk of injury.

## Conclusion

Under the study protocol, multilevel analyses of the body awareness construct will be performed to investigate its causal role in fall injuries during the physical activities that make up French Special Forces selection courses. Thus, the study will examine the influence of a body-centered prevention program. During these six, one-hour sessions, participants will be trained to actively pay attention to their own body signals, hopefully reducing the risk of fall injuries.

## Data Availability

Since the manuscript is a protocol paper there are no unpublished data. The original protocol (version 1.0 - 20210721) for this study is available on demand.

## Author Notes

The project is named ‘POSITION’, which is an acronym for a brief description in French: *Prévention de la blessure en milieu militaire par optimisation de la conscience corporelle* (Prevention of injuries in military settings based on body awareness enhancement). The opinions or assertions expressed herein are the private views of the authors, and are not to be considered as official or as reflecting the views of the French Military Health Service.

## Author Contributions

CV and AMD contributed equally to the work, including the conceptualization of the research question, and the design of the methodology. LO, CV and AMD contributed equally to the preparation of the paper. All the authors will take part in the execution of the study. They also approved the final version of the manuscript for submission.

## Dissemination

The study’s results will be disseminated at national and international conferences, and in peer-reviewed journals. The trial’s findings could also be made available to participants collectively. Each protocol modification will be submitted to competent authorities (if required) and communicated to all relevant parties, as specified under the French public health code.

## Data property

All data will remain the property of the sponsor. The final report will be prepared by all of the associated scientific investigators (as defined in the protocol).

## Informed Consent

Written informed consent will be obtained from all individuals included in the study.

## Roles and responsibilities

The study is sponsored by the *Direction centrale du service de santé des armées* (Site de Vincennes, 60, boulevard du général Martial Valin CS 21623, 75509 Paris Cedex 15). The coordinator, the clinical research associate coordinator, and the *Bureau de gestion de la recherche et de l’innovation* are in charge of logistics, notably: the quality of the collected data in the case report form - CRF, the creation of input masks, data computerization and quality control. Since the study does not involve the use of products or medications, and only represents a minor risk and constraint for participants, no data monitoring committee will be established.

## Supplementary Information files

Supplementary File 1: Details of the programs’ contents

### Sessions - Active Control Group

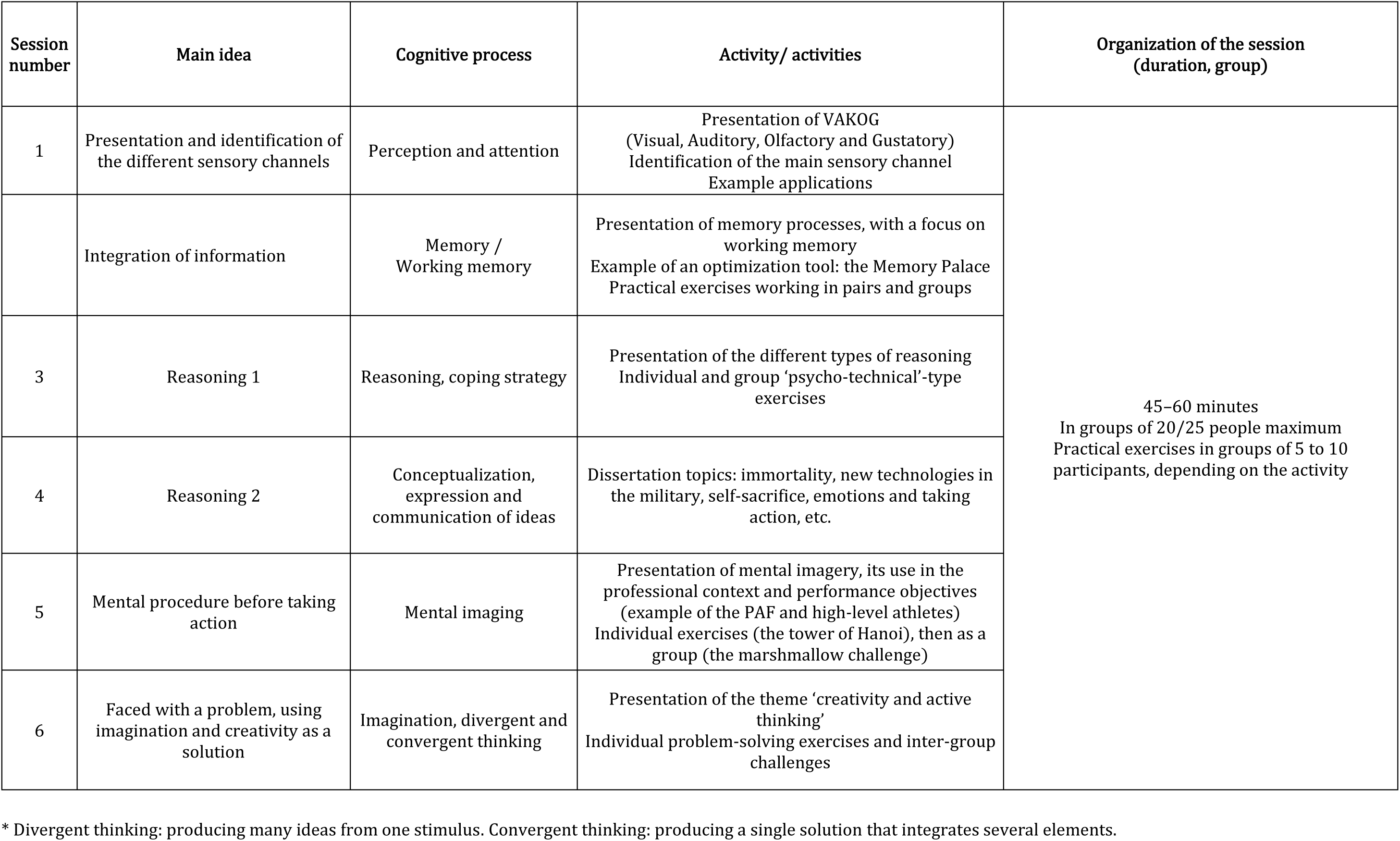

### Sessions - ORAF Group

| Session number | Theme | Techniques worked on | Content | Organization of the session (duration, group) |
| --- | --- | --- | --- | --- |
| 1 | ORAF Presentation - Breathing | Breathing - Relaxation | Presentation of the ORAF: philosophy, theory & application<br>Breathing: theory and practice | 45 to 60 minutes<br>In groups of up to 20 people |
| 2 | Sleep and Vigilance | Relaxing breathing and calm imagery | Sleep/Vigilance/Fatigue theory<br>Continual awareness of the whole body through relaxation (recovery and relaxation); using VAKOG to support an internal dialogue/relaxing imagery |  |
| 3 | Stress | The optimized activation signal (OAS) technique | Theoretical presentation of stress<br>Implementation of the OAS technique |  |
| 4 | Self-confidence, performance optimization | Relaxing breathing<br>Mental rehearsal | Mental rehearsal: presentation and practical exercise |  |
| 5 | Mindfulness and performance optimization | Geolocation (taking the environment into account)<br>Dynamic breathing | Practical geolocation exercise, optimized complete dynamization and immediate mental projection |  |
| 6 | Motivation and performance | Relaxation<br>Mental projection of goals (MPG) | Motivation theory + MPG<br>Individual relaxation exercise, followed by an MPG exercise |  |

